# Making the Exercise Experience More Pleasant: Effects of Open-Label Placebos on Affective Responses to Exercise Induced through Verbal Suggestions

**DOI:** 10.1101/2024.11.06.24316473

**Authors:** QIU Yue, YUN Dong-Ting, LIU Jiao, MAO Zhi-Xiong

## Abstract

**Objective:** By manipulating psychological factors—such as fostering positive expectations about exercise outcomes through verbal suggestions—it is possible to induce placebo effects without the use of traditional placebos, such as inert substances (closed-label placebos, CLPs). This can be achieved even when individuals are aware they are receiving a placebo through verbal suggestions (open-label placebo, OLP). This proof-of-principle study investigated whether the effects of OLPs on affective responses to exercise can be induced solely through verbal suggestions.

**Methods:** Eighty-nine healthy volunteers were randomized into three groups during a 30-minute session of moderate-intensity running. The first experimental group was informed that the running session would enhance their mood (CLP: positive verbal suggestion), the second experimental group was educated on the concept of an OLP and its anticipated effects during the running session (OLP: positive verbal suggestion), and the third control group received unrelated information about the study (no verbal suggestion). The primary study outcomes were self-reported affective valence during exercise and postexercise enjoyment. In addition, anticipated affect and remembered affect were assessed. Resting-state functional connectivity (RSFC) in the prefrontal cortex was objectively measured via functional near-infrared spectroscopy (fNIRS).

**Results:** Compared with the control group, both experimental groups presented increased affective valence during exercise, postexercise enjoyment, and anticipated affect, along with lower RSFC in the right prefrontal cortex, and the OLP intervention had a greater effect on remembered affect.

**Conclusions:** The results illustrate a potential role for OLPs in inducing affective response to exercise and suggest that further study of verbal suggestions through an extensive explanation of placebo effects might be promising for practice.

## Introduction

Despite compelling scientific evidence indicating the beneficial effects of both acute and chronic exercise on affective responses (1,2), a lingering concern regarding the extent to which these benefits can be attributed to the placebo effect remains (3,4). The placebo effect represents a favorable outcome stemming from an individual’s expected and/or learned response to a treatment or situation (3,5). The findings of relevant meta-analyses indicate that the actual impact of exercise training on psychological outcomes is approximately half of what has been previously reported (the mean effect size of the placebo was 0.20, and the observed effect of exercise training was 0.37) (6). Thus, this sparked interest in placebo effects and led to an increase in research activities. By manipulating psychological factors—such as fostering positive expectations about the effects of exercise—the placebo effect can be induced, thereby enhancing the outcomes of exercise (7).

The term ‘placebo’ is commonly defined as an inert substance or procedure (3,5). However, over the past 50 years, interviewees have embraced a broad definition of placebos. Placebos come in various forms and have evolved from tangible inert substances or therapies to intangible verbal suggestions (3,7,8). For example, experimental studies have further demonstrated that placebo effects on exercise outcomes can be induced by providing verbal suggestions that exercise is able to boost one’s mood (4,9,10,11,12,13). Placebo effects can be established by an individual’s belief in the effectiveness and positive outcomes of exercise (7). The main working mechanisms of placebo effects include associative learning processes, such as conditioning, and the adjustment of expectations, such as by providing positive information regarding treatment outcomes by means of verbal suggestions (14,15,16).

Conventional placebos are typically administered by experimenters in a concealed manner (17), and the concealment of their administration has long been considered a fundamental component in the elicitation of the placebo effect (18). For example, most studies on the placebo effect employ an experimental approach, eliciting the effect by providing overstated or deceptive information about the specific exercise regimen offered (19). This overstated or deceptive component complicates the potential utilization of placebo effects in clinical practice, considering that the omission of treatment information and provision of deceptive information are unethical (3). Considering these concerns, Open-label placebos (OLPs) have emerged as a potential solution. OLPs are administered without deception. The administration of OLPs involves openly informing participants about the nature of the placebo intervention, including the conceptual framework and scientific principles. Accumulating evidence has revealed the efficacy of OLPs for the treatment of pain (20), depression (21), and anxiety (16,22) and the improvement of psychological well-being (23). Most studies on OLP treatment have reported medium-to-large effect sizes (24,25), which are comparable to the effect sizes reported in studies in which patients were not informed about receiving an inert substance (CLP group).

The mechanisms through which OLP effects may arise remain unclear. Previous OLP studies have incorporated various components, including the administration of inert pills and explanations about the effectiveness of placebos and their mechanisms (14, 24,25,26). This complexity makes it challenging to discern which components contribute to the OLP effect and to what extent. It is uncertain whether merely providing positive explanations (e.g., verbal suggestions) is sufficient to induce an OLP effect by altering expectations of exercise outcomes and influencing emotional responses to exercise stimuli. If the induction of these effects is shown to be possible, it could enhance practical applications, such as the optimization of adherence to existing exercise prescriptions through improved patient–provider communication.

Given the subjective nature of exercise-induced affective responses, it is important to investigate whether placebo effects can yield authentic and advantageous physiological changes or if they are merely outcomes of response biases induced by participants’ conformity to experimenter expectations. This inquiry is further highlighted in the context of the placebo effect; in the domain of exercise, the prefrontal cortex (PFC), a brain region implicated in cognition and mood regulation, is partially involved in running and forthcoming affective responses (27,28). Activation of the left PFC has been consistently documented after short-duration aerobic exercise (29,30,31), underscoring a nuanced interplay between frontal lobe lateralization and affective responses as well as motivation. Notably, activation of the left PFC appears to be associated with positive affect and approach motivation, whereas activation of the right PFC is correlated with negative affect and avoidance motivation (32,33). On the basis of these properties of the placebo effect and running, the neural and physiological changes associated with exercise and the placebo effect are similar. We hypothesized that running combined with the placebo effect has the potential to enhance mood with broad prefrontal activation and that activation is evident in the left PFC region.

### The present study

For the first time, the present study aimed to investigate whether positive verbal suggestions about exercise outcomes in response to exercise-induced affective responses, without combining them with an inert treatment, can induce positive outcome expectations through an OLP design. Additionally, we investigated neurophysiological changes by using functional near-infrared spectroscopy (fNIRS) to measure the activation levels of the PFC.

Self-reported affective valence during exercise and postexercise enjoyment were the primary study outcomes, and anticipated affect, remembered affect and resting-state functional connectivity (RSFC) in the PFC were the secondary outcomes. We hypothesized the following: (1) Compared with the participants in a control group who received no verbal suggestions, participants in the OLP group (positive verbal suggestions) and the CLP group (positive verbal suggestions) would report more positive affective responses after short-duration aerobic exercise. Given the exploratory nature of studies that simultaneously compare the effects of OLP and conventional placebo interventions, there is no directional hypothesis regarding the superiority of either intervention. (2) The improved affective response induced by the OLP and CLP interventions would be reflected by the engagement of a network of brain regions previously associated with exercise and placebo effects.

## Methods

### Study Design

A mixed experimental randomized controlled trial design was used. **Affective response during exercise**: For this variable, a 3 (group: CLP vs. OLP vs. control) × 6 (time: 5 minutes, 10 min, 15 minutes, 20 minutes, 25 minutes, 30 minutes) mixed experimental design was used; **RSFC after exercise**: For this variable, a 3 (group: CLP vs. OLP vs. control) × 2 (time: pretest vs. posttest) mixed experimental design was used. Group was a between-group variable, whereas time was a within-group variable. The randomization sequence was generated via an online random number generator (www.random.org, Dublin, Ireland). The laboratory session was overseen by two experimenters, and the experimental assistant responsible for group allocation and the delivery of verbal suggestions to the participants was distinct from the individual in charge of overseeing the study outcomes and conducting the exercise test.

## Data Availability Statement

The data can be obtained by corresponding author.

## Ethics Statement

This study was approved by the Institutional Review Board of the School of Psychology at Beijing Sports University in China (ethics approval number 20211005).

## Participants

A priori power calculations were conducted with G*Power 3.1 to determine the sample size. According to the following values, the total sample size was calculated to be 36: during exercise: *f* = 0.25, α = 0.05, 1 − β = 0.8, number of groups = 3, measurement times = 6, and within-between interaction. According to the following values, the total sample size was calculated to be 66: after exercise, *f* = 0.25, α = 0.05, 1 − β = 0.8, number of groups = 3, measurement times = 2, and within-between interaction. A total of 89 eligible participants (*M*age = 19.26 ± 2.07 years), including 16 males and 73 females, were recruited. Participants who had previously taken part in a similar experiment, possessed prior knowledge of the study’s objectives, or exhibited any contraindications to exercise, as indicated by the Physical Activity Readiness Questionnaire (PAR-Q) (34), were excluded from the study.

## Materials and Measures

### Exercise Prescription

The exercise intensity was determined via the heart rate reserve (HRR). The HRR was calculated by subtracting the resting heart rate (HRrest) (measured after a 5-minute period of rest in a seated position) from the maximum heart rate (HRmax= 207–0.7 * age) (with the formula: (HRmax – HRrest) * (xx%) + HRrest). The heart rate range for moderate-intensity exercise was set at 60–69% of the HRR, with a specific target heart rate of 65%, which is considered roughly indicative of moderate-intensity exercise on a treadmill according to the American College of Sports Medicine (ACSM) guidelines (35). The aerobic exercise duration consisted of a 30-minute session, which included a 5-minute warm-up, 20 minutes of exercise at the target intensity, and a 5-minute cool-down period.

### Verbal Suggestions Intervention

The participants were informed prior to the session that the study aimed to investigate individual differences in the experience of exercise. Upon arrival at the laboratory, the following general instructions were given to all participants: “During the test, we will conduct a 30-minute moderate-intensity treadmill run, during which your heart rate will be monitored to maintain exercise intensity. Please pay attention to your affective experiences throughout the running session.” (1) In the CLP group, the participants were given the following verbal suggestion (e.g., “Notably, previous research has shown that 95% of healthy individuals experience a positive affective response during moderate-intensity running. They reported experiencing a sense of pleasure and satisfaction, accompanied by heightened levels of energy and vitality”). This finding highlights the potential psychological benefits associated with engaging in such exercise routines. (2) In the OLP group, building on the framework of verbal suggestions as CLPs, we incorporated a direct explanation of the concept of placebos for the participants, along with an overview of the scientific principles that underlie their efficacy. The specific intervention content strictly followed the relevant guidelines (conventional OLP disclosures) (14,15) (e.g., “Previous research has shown that 95% of healthy individuals experience a positive affective response while running. Furthermore, studies have indicated that expectations significantly influence the exercise experience. For example, by providing information about what to expect from running, such as this session, I just told you that exercise sessions often result in a more favorable emotional state. Research has demonstrated that such suggestions can enhance individuals’ emotional well-being, even when they are aware of receiving this suggestion.” (3) In the control group, no verbal suggestions were provided to the participants.

### Manipulation Check

Building upon the previous research (9,36), a manipulation check was conducted in this study to measure the participants’ levels of trust in the verbal suggestion intervention. Question 1 was “How do you think your affective response will be affected during the upcoming 20-minute moderate-intensity run? Please indicate your assessment using a bipolar Likert scale, where -7 indicates “a significant negative impact,” 0 represents “no effect,” and 7 denotes “a considerable positive influence.”. Question 2 was “On a scale from -3 (completely inconsistent) to 3 (fully consistent), please rate the extent to which you believe the information presented in this study aligns with your past experiences”. Participants who received a score equal to or below 0 on Question 2 were excluded from this study.

### Primary outcome measure: self-reported affective response to exercise

**Outcome Measure During Exercise** The *Feeling Scale* (FS) (37) was used in this study. Participants are provided the following statement: “Use the following numbers to show how you feel at this time.” The item is scored on an 11-point Likert scale ranging from −5 (extremely bad) to 5 (extremely good), with 0 being neither good nor bad. The scale range was explained and anchored; the participants had the opportunity to give practice ratings so that they were fully familiarized with the FS. The FS was completed every 5 minutes during the running session (e.g., 5 minutes, 10 min, 15 minutes, etc.) ***Outcome Measure Postexercise.*** *The Physical Activity Enjoyment Scale* (PACES) (38) was used in this study. The PACES is an 18-item measure, and respondents indicate how they feel following exercise on a scale from “1 = boring” to “7 = enjoy”. The scale was designed to be used immediately after exercise; thus, it was administered only once following the exercise session. The internal consistency alpha coefficient was 0.87.

### Secondary Outcome Measure: Remembered affect and anticipated affect

Adapted from the self-designed single-item questions utilized by Kwan et al. (11), the prompt for remembered affect was “Reflect on your emotional state immediately following the running session: How do you recall your feelings on a scale from -100 (extremely unpleasant) to 100 (extremely pleasant). The anticipated affect instructions were as follows: How do you expect your affective response to be influenced during your next run? Please provide a rating on a scale from -7 (indicating an extremely negative impact) to 7 (indicating an extremely positive impact).

### Secondary Outcome Measure: Resting-state functional connectivity (RSFC) in the prefrontal cortex

A prototype modular, high-density DOT system known as “LUMO”, developed by the University of London (UCL)-affiliated Gowerlabs Ltd., was employed in this research (39,40). The device integrates transmitters and receivers into a hexagonal tile, with each tile containing 3 sources and 4 detectors. For this study, 7 tiles were selected, resulting in 42 channels with distances ranging from 10 mm to 40 mm. The tiles were arranged in a double rhombus pattern and were symmetrically distributed on both sides of the forehead (Figure 1). Near-infrared light with wavelengths of 735 nm and 850 nm was utilized, with a sampling rate of 10 Hz/m. The following channels cover the following six regions of interest (ROIs): the right dorsolateral prefrontal cortex (rDLPFC), which is covered by channels CH1–CH6; the right frontopolar area (rLFPA), which is covered by channels CH27–CH29 and CH31–CH38; the right orbitofrontal cortex (rOA), which is covered by channels CH39–CH42; the left dorsolateral prefrontal cortex (lDLPFC), which is covered by channels CH7–CH12; the left frontopolar area (lFPA), which is covered by channels CH15, CH16, CH19–CH26, and CH30; and the left orbitofrontal cortex (lOA), which is covered by channels CH13, CH14, CH17, and CH18.

**Figure 1.**
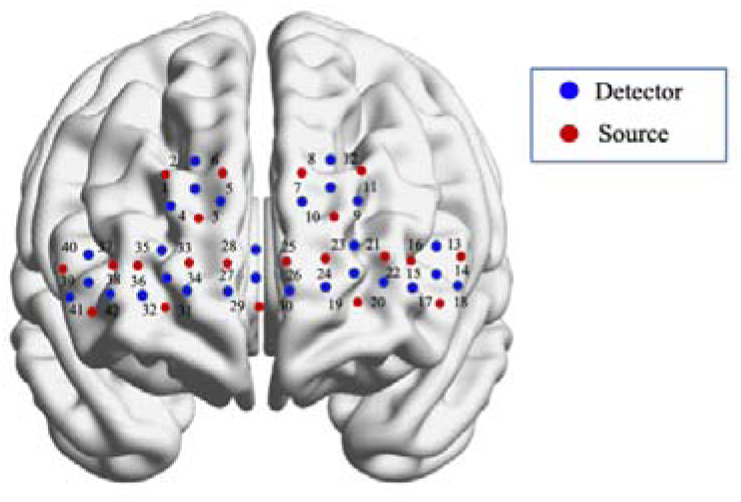
Seven tiles over the prefrontal cortex yielded 21 sources (red points) and 28 detectors (blue points).

**Figure 2.**
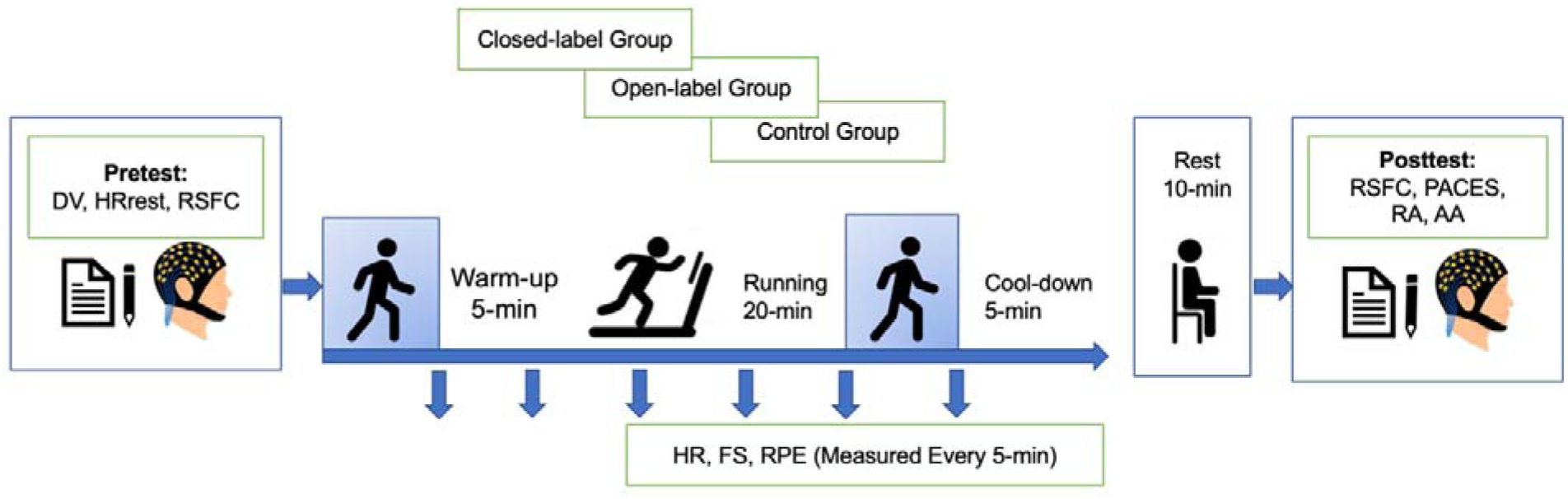
Experimental flow chart Note: DV: demographic variables; RSFC: resting-state functional connectivity; HR: heart rate; FS: feeling scale; RPE: rating of perceived exertion scale; PACES: The Physical Activity Enjoyment Scale; RA: remembered affect; AA: anticipated affect.

This study assessed the strength of RSFC features in the PFC before and after exercise. During resting-state data acquisition, the participants were instructed to fixate on a “+”, remain still, avoid head movements, and maintain wakefulness for a continuous period of 10 minutes. The resting-state data collection included preexercise and postexercise measurements, with the postexercise measurement conducted 15 minutes after the exercise session. The fNIRS indicators encompassed the calculation of the mean resting-state whole-brain functional connectivity (RSFC) using the oxygenated hemoglobin concentration (HbO) signal, along with the assessment of RSFC values among 42 channels. A stronger RSFC signifies an increased simultaneous enhancement in the signal strength of the two-time series between the channels, indicating heightened coordinated activity in the respective brain region.

### Controlled variable: Physical activity

The Godin Leisure-Time Exercise Questionnaire (GLTEQ) (41) was used in this study. This scale assesses the frequency of exercise performed at different intensities within the past week, including low-, moderate- and high-intensity exercise, with each session lasting for more than 20 minutes. The GLTEQ comprises three items and is scored as follows: (frequency of low-intensity exercise × 3) + (frequency of moderate-intensity exercise × 5) + (frequency of high-intensity exercise × 9). A higher total score indicates a higher level of physical activity.

### Heart Rate

The participants’ heart rates (HRs) were continuously monitored during the exercise sessions via Polar heart rate sensors (chest straps) to ensure that they maintained moderate-intensity exercise. This sensor is a reliable and accurate tool for real-time HR measurement in adults during exercise and shows a strong correlation with other devices, such as simultaneous electrocardiograms (ECGs) (42). The collected data were extracted for analysis via PolarFlow software. **Perceived Exertion**: The Rating of Perceived Exertion (RPE) scale (43) was utilized to assess whether participants maintained moderate-intensity exercise during the sessions. The participants were asked “How is your perceived exertion right now?” and responded on a scale ranging from 6 (no exertion at all, such as when sitting or resting) to 20 (maximum exertion). This scale was displayed in color on a hardboard (70 cm × 40 cm) mounted on the wall in front of the participants. Before the warm-up, the participants were informed that the experimenter would check their status every five minutes, and they indicated their response by pointing to the appropriate number on the board. The RPE scale is a validated tool for determining exercise intensity, with an RPE of 11–13 indicating moderate intensity (44).

### Procedure

#### Stage 1: Preparation

1. After providing informed consent, the participants completed a set of questionnaires that collected demographic information.
2. The participants were then required to wear a Polar H10 heart rate monitor to measure their HRR and to determine the appropriate exercise intensity.
3. A 10-minute pretest of resting-state frontal lobe functional connectivity was conducted via an fNIRS device.

#### Stage 2: Placebo and exercise intervention

1. The participants were randomly assigned to one of three groups, and each group of participants received the assigned induced expectancy intervention as planned; subsequently, a manipulation check was carried out.
2. Each participant was prescribed a 30-minute aerobic running session with the moderate intensity set according to the HRR. The HR of each participant was monitored throughout the exercise session via a Polar sports tester (Polar A300), while the FS was completed every five minutes (0–5 minutes, 5–10 minutes, 10–15 minutes, etc.).

#### Stage 3: End of the session

1. After the cool-down period, posttests were administered to measure changes in mood states and perceived affect via the PACES.
2. After 15 minutes of the exercise session, a 10-minute resting-state RSFC measurement was conducted. Damrongthai et al. (27) reported that all physiological variables returned to baseline levels within a span of 15 minutes. Hence, to mitigate potential signal contamination, we conducted fNIRS measurements 15 minutes postexercise.

### Statistical analyses

The questionnaire data were analyzed primarily via JASP 0.16.1.0. The normality of the scale data was assessed via the ShapiroLWilk test. To examine the effects of group and measurement time points on the outcomes, a repeated-measures analysis of variance (ANOVA) with Greenhouse–Geisser correction was conducted. In cases where a significant F-ratio was observed, LSD post hoc tests were performed for pairwise comparisons. Effect sizes were calculated as partial η_p_^2^ for significant differences in pairwise comparisons. Statistical significance was defined as *p* ≤ .05, and alpha levels were adjusted to .05.

Statistical analysis of the fNIRS data was performed with MATLAB 2021Ra software, the DOT-HUB toolbox, and the Homer2 FC-NIRS processing package BrainNet was used for conducting brain functional imaging. Prior to data analysis, several preprocessing steps were performed: (1) channel selection, wherein channels with a signalLtoLnoise ratio less than 12 were discarded; (2) motion artifact removal, where data with signal amplitude changes exceeding 20 standard deviations within a 2-second interval were eliminated; (3) filtering, which involved setting the highLpass filter at 0.01 Hz and the lowLpass filter at 0.5 Hz; and (4) conversion of the optical density to the hemoglobin concentration. Finally, functional RSFC was calculated via the preprocessed data. With the FC-NIRS toolkit, the RSFC of the PFC was computed via overall HbO signals. This involved calculating the average RSFC value across all channels, as well as the mean RSFC value for each pairwise channel combination, resulting in a total of 861 such combinations derived from 42 channels. To assess RSFC differences, paired-sample t tests were employed for within-group pre- and posttest comparisons, whereas analysis of covariance (using pretest RSFC as a covariate) was utilized for between-group comparisons. To mitigate potential false positive results arising from multiple comparisons, the false discovery rate (FDR) method was implemented for correction.

## Results

### Sample characteristics

The demographics and sample characteristics of the 89 participants who completed the experimental session are shown in Table 1.

**Table1.**
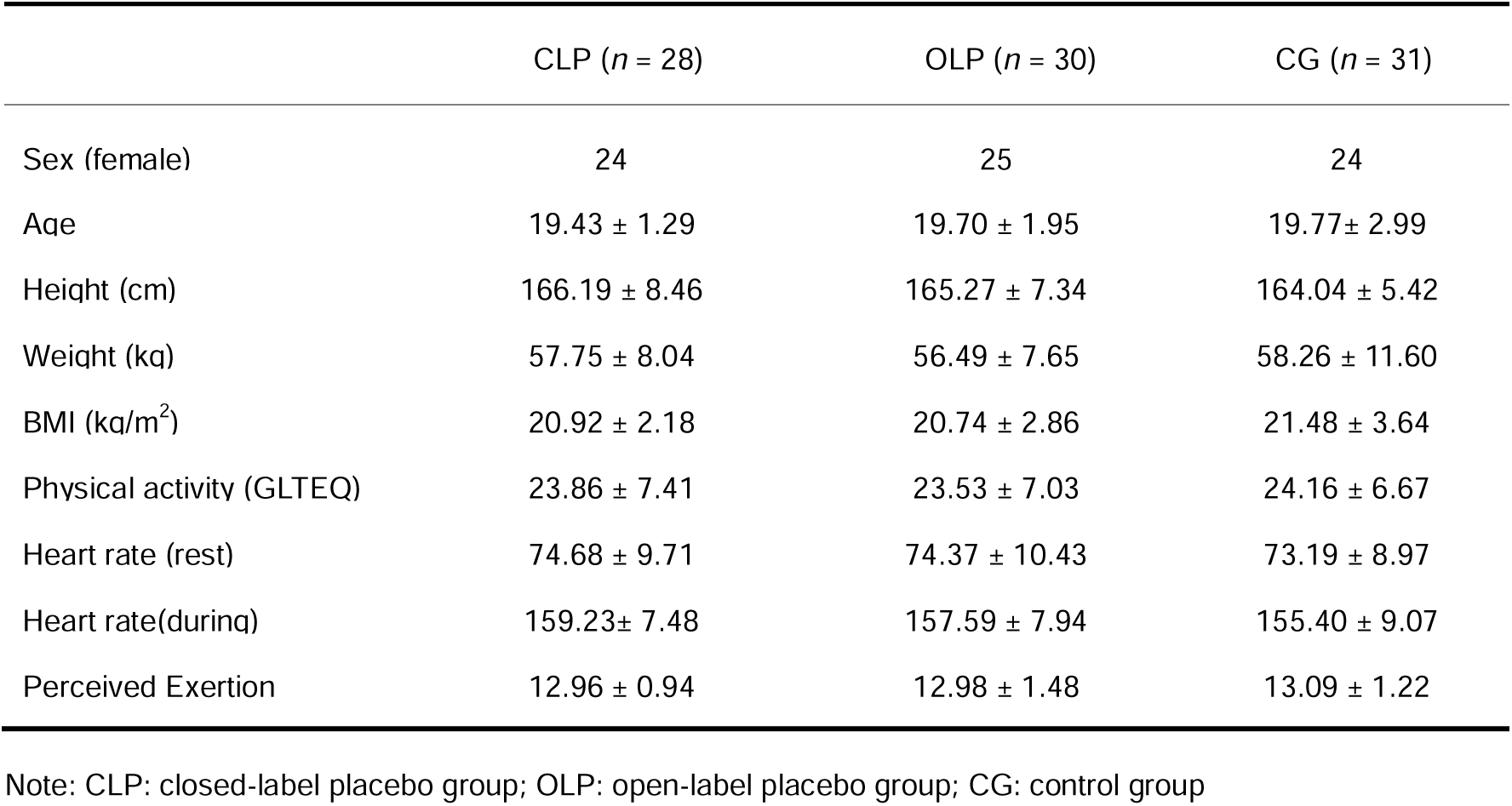
Mean and Standard Deviation (SD) for demographics variables and sample characteristics.

|  | CLP (*n* = 28) | OLP (*n* = 30) | CG (*n* = 31) |
| --- | --- | --- | --- |
| Sex (female) | 24 | 25 | 24 |
| Age | 19.43 ± 1.29 | 19.70 ± 1.95 | 19.77± 2.99 |
| Height (cm) | 166.19 ± 8.46 | 165.27 ± 7.34 | 164.04 ± 5.42 |
| Weight (kg) | 57.75 ± 8.04 | 56.49 ± 7.65 | 58.26 ± 11.60 |
| BMI (kg/m<sup>2</sup>) | 20.92 ± 2.18 | 20.74 ± 2.86 | 21.48 ± 3.64 |
| Physical activity (GLTEQ) | 23.86 ± 7.41 | 23.53 ± 7.03 | 24.16 ± 6.67 |
| Heart rate (rest) | 74.68 ± 9.71 | 74.37 ± 10.43 | 73.19 ± 8.97 |
| Heart rate(during) | 159.23± 7.48 | 157.59 ± 7.94 | 155.40 ± 9.07 |
| Perceived Exertion | 12.96 ± 0.94 | 12.98 ± 1.48 | 13.09 ± 1.22 |

Note: CLP: closed-label placebo group; OLP: open-label placebo group; CG: control group

### Manipulation Check Results

Regarding the manipulation check for Question 1, ANOVA yielded significant results, indicating a significant main effect of group *(F* (2, 85) = 15.16, *p* < .001, η_p_^2^ = 0.25). Subsequent pairwise comparisons revealed that both the CLP (*M* = 5.14, *SD* = 1.56) and the OLP groups (*M* = 4.41, *SD* = 1.42) demonstrated significantly greater positive anticipated outcomes than did the control group (*M* = 2.77, *SD* = 2.03; *t* = -5.34, *p* < .001, *d* =1.39; *t* =-3.73, *p* < .001, *d* = 0.96). In the manipulation check for Question 2, two participants were excluded from the analysis because they expressed disbelief in the information provided about the placebo. These findings support the validity of the experimental procedures employed in this research.

### Verbal suggestions on affective response during exercise

Repeated measures ANOVA was conducted, which revealed that the main effect of group was significant (*F*(2, 85) = 11.51, *p* < .001, η_p_^2^ = .21). Post hoc tests revealed that both the CLP group and the OLP group had greater affective valence than did the control group (*t* = 3.46, *p* = .002, *d* = 0.78; *t* = 4.59, *p* < .001, *d* = 1.02). The main effect of measurement time was significant (*F*(3.24, 275.71) = 33.46, *p* < .000, η_P_^2^ = .28). This finding indicates that affective valence changed (became more positive) as the exercise session progressed (*t* > 3.28, *p* < .05, *d* > 0.29). The interaction effect between group and time was not significant (*F*(6.49, 275.71) = 1.41, *p* = .21, η_p_^2^ = .03), which suggests that the trend of the change in affective valence over time was similar across the three groups, and the measurement changes in affective valence are depicted in Figure 3.

**Figure3.**
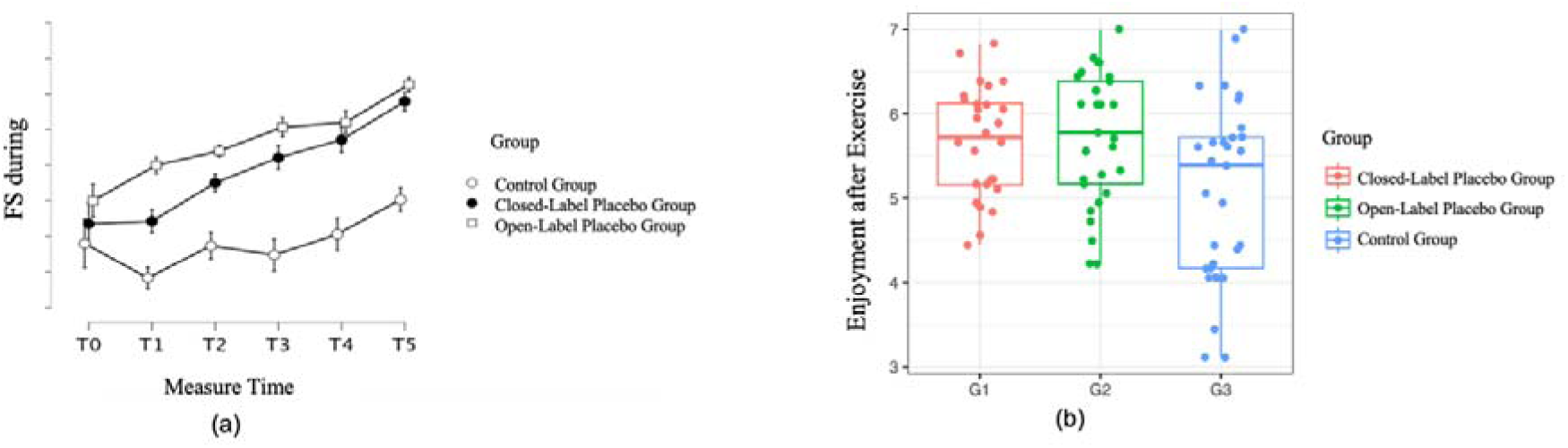
Figure(a): The trend of affective valence changes during exercise of in each group, as measured by the feeling scale; Figure(b): The trend of enjoyment changes after exercise in each group; enjoyment measured using the PACES.

### Verbal suggestions on affective response after exercise

One-way ANOVA was conducted to examine the impact of group on postexercise enjoyment, as measured by the PACES. The results revealed a significant main effect of group on postexercise enjoyment (*F*(2, 85) = 4.68, *p* = 0.01, η_p_^2^ = 0.10). Specifically, the participants in the OLP group reported significantly greater levels of enjoyment (*M* = 5.72, *SD* = 0.78) than did those in the control group (*M* = 5.06, *SD* = 1.06) (*t* = 2.86, *p* = 0.02, *d* = .74). Moreover, the participants in the CLP group (*M* = 5.60, *SD* = 0.77) also reported significantly higher enjoyment levels than did those in the control group (*t* = 2.23, *p* = 0.047, *d* = 0.61); please refer to Figure 4 for a visual representation of these findings.

**Figure 4.**
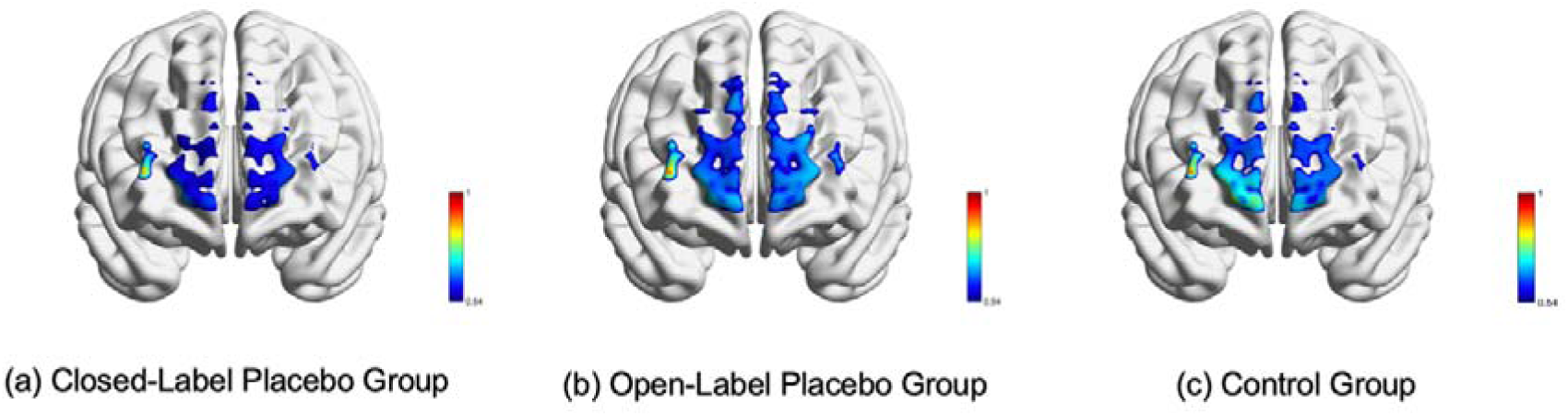
Neuroimaging results Figures (a), (b), and (c) depict the scalp-based mapping of Pearson correlation coefficients between the CH37 seed point channel and the time series of each voxel in the entire brain. These maps provide insights into the variations in functional connectivity strength within the frontal lobe across different groups.

ANOVA was conducted to examine the effects of different groups on the remembered affect and anticipated affect. The results revealed a significant main effect of group on the remembered affect (*F*(2, 85) = 3.83, *p* = .039, η_p_^2^ = 0.069). Post hoc analyses indicated that the participants in the OLP group exhibited more positive remembered affect than did those in the control group (*t* = 2.49, *p* = .038, *d* = 0.64). In terms of anticipated affect, a significant main effect of group was observed (*F*(2, 85) = 3.96, *p* = .023, η_p_^2^ = 0.09). Further post hoc analyses revealed that both the CLP and OLP groups reported more positive anticipated affect than did the control group (*t* = 2.10, *p* = 0.04, *d* = 0.55; *t* = 2.65, *p* = .026, *d* = 0.69), with no significant difference between the two placebo groups. The descriptive statistical results are shown in Table 2.

### Verbal suggestions for fNIRS parameters

After the data quality evaluation, 89 participants were selected for data processing and analysis. **Within-group differences in RSFC:** The average strength of connectivity across 861 edges, represented by correlation coefficients between each pair of channels, was used as an indicator of the mean RSFC of the PFC. To explore the differences in RSFC between groups before and after exercise, independent samples t tests were employed. The results revealed a significant increase in the total mean RSFC of the PFC after exercise in all groups compared with the pretest value *(t values* > 8.11, *p values* < 0.001; *d values* > 0.28). **Between-group differences in RSFC:** One-way analysis of covariance (ANCOVA) was performed, with group as the between-group independent variable, the total mean value of RSFC as the dependent variable, and pretest RSFC as the covariate. The data satisfied the assumptions necessary for conducting ANCOVA. The results revealed a nonsignificant main effect of group (*F* (2, 85) = 1.3, *p* = .27, η_p_^2^ = .01), indicating that verbal suggestions did not significantly influence the overall mean value of RSFC. ANCOVA was subsequently conducted using the 861-connectivity channel “edges” as the dependent variable. The results revealed significant differences across 21 edges after FDR correction for multiple comparisons (*F values* > 3.35, *p values* < 0.041). The significant edges included CH1–CH2, CH1–CH10, CH9–CH34, CH17–CH40, CH18–CH39, CH18–CH40, CH19–CH22, CH23–CH40, CH33–CH37, CH33–CH40, CH35–CH37, CH37–CH38, CH37–CH40, CH37–CH42, CH38–CH40, CH39–CH40, CH39–CH41, CH33–CH40, CH40–CH41, CH40–CH42, and CH41–CH42. Post hoc tests revealed that the control group had significantly greater RSFC in the aforementioned channels than both the CLP group and the OLP group did. These channels were located mainly in the right area. However, there were no significant differences between the CLP and the OLP verbal suggestion interventions. Among the significantly different edges, CH37 had the highest number of connections. Consequently, CH37 was chosen as the seed point, and the Pearson correlation coefficient was computed between its time series and the time series of each voxel in the frontal lobe to perform functional brain imaging (Figure 5).

## Discussion

The aim of this study was to investigate the potential effects of placebos involving verbal suggestions on affective response during and after short-term aerobic exercise, as well as to explore the related neurophysiological changes. Through behavioral data analysis, the results of this study support our hypothesis that both CLPs and OLPs can significantly improve positive affective response during exercise, with a large effect size. Additionally, these interventions significantly increased postexercise feelings of pleasure and vigor, with a medium to large effect size. This finding provides evidence that the placebo effect can be present in nondeceptive conditions, which is in line with previous research in the field of clinical medicine (14,45). This is attributed to the proactive nature of individuals, who do not passively await outcomes but actively select and influence them (46) . When individuals anticipate positive affective responses following exercise, they proactively prepare for these outcomes or actively modify the likelihood of occurrence. In addition, in light of inconclusive evidence regarding the emotional benefits of exercise, the pervasive influence of social media has cultivated a widespread belief that exercise yields positive experiences. This familiarity with the belief facilitates the acceptance of this expectation, thereby contributing to the generation of a placebo effect.

However, in comparison with the CLP group, only the OLP group had a significant effect on enhancing the remembered affect. In the realm of affective memory retrieval, it is crucial to acknowledge the active and reconstructive nature of the memory process. Rather than being stored as static textual records for mere playback, memories of past events and their associated emotional states undergo a dynamic process of reconstruction (47). This reconstruction process is intricately influenced by a diverse range of factors, encompassing an individual’s current emotional state and expectations (48). Consequently, the amplification of affective responses following exercise can elicit more positive remembered affect, thereby influencing subsequent affective experiences (49). These findings suggest that OLPs may have advantages in long-term exercise interventions.

This could be attributed to the fact that OLP interventions evoke a greater sense of uncertainty, curiosity, and cognitive dissonance than CLP interventions do. As a result, they enhance participant motivation and facilitate increased conscious introspection and self-evaluation (45). The impact of OLPs highlights the importance of individuals maintaining an open-minded and receptive attitude, being willing to give exercise a try, increasing their level of self-involvement and engagement, and experiencing a heightened subjective sense of agency (45,50).

This study employed fNIRS technology to investigate changes in blood oxygen signal levels in the PFC of the brain. Following moderate-intensity acute aerobic exercise, a significant increase in RSFC was observed among all participants, accompanied by greater postexercise enjoyment. These findings align with those of previous studies indicating that exercise promotes enhanced functional connectivity in networks involved in affective processing (51), providing further evidence of neuroplasticity in the brain. However, the effect of the placebo intervention involving verbal suggestions did not result in additional enhancement of neuroplasticity. Nonetheless, after short-term moderate-intensity aerobic exercise, both the CLP and the OLP groups displayed more enjoyment following exercise than did the control group. Furthermore, a significant decrease in RSFC was observed in the R-PFC. However, no significant differences were found in terms of RSFC in the left prefrontal cortex among the three groups, which is in line with the findings reported by Damrongthai et al (27). After a 10-minute moderate-intensity run, followed by a 15-minute rest period, all areas of the PFC, except for the lDLPFC, were activated. This finding underscores the importance of carefully timing measurements following exercise in research studies. In low-intensity exercise below the ventilatory threshold (VT), frontal asymmetry predicts immediate postexercise emotions at a rate of 12.8%. However, this predictive value increases to 15.8% when it is measured 20 minutes after the completion of exercise. These results provide further support for the valence asymmetry hypothesis with respect to the PFC (32). The valence asymmetry hypothesis posits that affect associated with avoidance motivations, such as disgust and fear, is characterized by greater activation in the R-PFC, whereas affect related to approach motivations, such as joy and satisfaction, is characterized by greater activation in the left PFC (33). As highlighted in a comprehensive systematic review conducted by Haehl et al. (2022)(29), converging evidence from functional magnetic resonance imaging (fMRI), near-infrared spectroscopy (NIRS), and electroencephalography (EEG) studies consistently demonstrated distinctive activation patterns within the PFC during tasks involving negative emotional stimuli. The valence asymmetry effect was observed through heightened activation in the R-PFC during efforts to inhibit and regulate negative responses that arise during exercise, such as fatigue and pain (28). In this study, no significant differences were found in RSFC in the left PFC among the groups. However, the control group exhibited significant activation in the right frontal lobe. This finding suggests that individuals in the control group were more motivated to avoid acute aerobic exercise and were more inclined to experience and express negative emotions. Furthermore, a bidirectional relationship was observed between bodily sensations and activity in the R-PFC. Specifically, higher levels of pain or discomfort are associated with increased activity in the right PFC, which, in turn, contributes to the downregulation of pain or discomfort (30,31). Hence, it is hypothesized that the placebo effect operates by attenuating negative affective experiences and buffering the behavioral inhibition system, leading to heightened affective responses after exercise. This effect may be mediated through a decrease in RSFC within the right frontal lobe.

## Limitations and future research directions

First, the activity patterns of the PFC during exercise were not monitored via fNIRS. While the placebo effect has been found to enhance affective responses during exercise, the specific interaction between the placebo effect and the PFC, as well as the underlying neural mechanisms involved, remains unclear. In future studies, it will be crucial to differentiate affective responses during exercise from those after exercise. Additionally, researchers should explore the neurophysiological changes associated with the placebo effect at different time points and investigate the underlying neural mechanisms triggered by expectancy.

Second, in all research involving placebo effects from verbal suggestions, reporting bias cannot be discounted, as participants may alter their responses on the basis of explicit expectations provided.

Furthermore, the specific mechanisms underlying the effects of CLPs and OLPs involving verbal suggestions have not been thoroughly investigated. Compared with CLPs, OLPs are associated with heightened curiosity, which, in turn, drives internal motivation and reduces individuals’ prediction errors and information gaps, ultimately diminishing individuals’ uncertainty about their surroundings (45). Therefore, further research should continue to explore this idea in greater depth and investigate the potential role of curiosity as a mediator of nondeceptive placebo effects.

Finally, the potential of enhanced affective responses following placebo interventions to promote subsequent exercise behavior has not been thoroughly explored. Given the harmless nature and cost-effectiveness of placebo interventions involving verbal suggestions, they hold significant practical value for exercise practitioners and researchers. Therefore, future research should incorporate long-term placebo interventions to comprehensively investigate their effects on exercise behavior.

## Practical implications

There is growing support for the promotion of placebos involving verbal suggestions and their recommended use to enhance affective responses to exercise. First, these interventions have been found to yield more consistent and reliable effects, with higher levels of compliance observed. Second, by fully considering participants’ rights to informed consent, OLP interventions address the ethical controversies surrounding placebo use, thus alleviating concerns about trust erosion and potential negative outcomes associated with deception (15)(Locher et al., 2021).

## Conclusion

Both the CLP and OLP (verbal suggestions) demonstrated positive effects on affective responses. However, the effects of the OLP intervention appeared to be more robust for remembered affect. The placebo effects on affective responses following exercise were modulated by asymmetrical patterns of prefrontal activity, whereby reduced functional connectivity in the R-PFC inhibited negative affective responses.

## Reference

1. Chekroud SR, Gueorguieva R, Zheutlin AB, Paulus M, Krumholz HM, Krystal JH, et al. Association between physical exercise and mental health in 1·2 million individuals in the USA between 2011 and 2015: a cross-sectional study. Lancet Psychiatry. 2018 Sep;5(9):739–46.

2. Ding D, Mutrie N, Bauman A, Pratt M, Hallal PRC, Powell KE. Physical activity guidelines 2020: comprehensive and inclusive recommendations to activate populations. Lancet. 2020 Nov 25;396(10265):1780–2.

3. Beedie C, Benedetti F, Barbiani D, Camerone E, Cohen E, Coleman D, et al. Consensus statement on placebo effects in sports and exercise: The need for conceptual clarity, methodological rigour, and the elucidation of neurobiological mechanisms. Eur J Sport Sci. 2018 Aug 16;18(10):1383–9.

4. Lindheimer JB, O’Connor PJ, McCully KK, Dishman RK. The Effect of Light-Intensity Cycling on Mood and Working Memory in Response to a Randomized, Placebo-Controlled Design. Psychosom Med. 2016 Aug 23;79(2):243–53.

5. Enck P, Bingel U, Schedlowski M, Rief W. The placebo response in medicine: minimize, maximize or personalize? Nat Rev Drug Discov. 2013 Mar 1;12(3):191–204.

6. Lindheimer JB, O’Connor PJ, Dishman RK. Quantifying the placebo effect in psychological outcomes of exercise training: a meta-analysis of randomized trials. Sports Med. 2015 Mar 11;45(5):693–711.

7. Qiu Y, Mao Z, Yun D. Can the add□on placebo effect augment the physical and mental health outcomes of exercise? A meta□analysis. Applied Psych Health & Well. 2021 Nov 8;14(2):483–98.

8. Petrie KJ, Rief W. Psychobiological Mechanisms of Placebo and Nocebo Effects: Pathways to Improve Treatments and Reduce Side Effects. Annu Rev Psychol. 2018 Aug 15;70(1):599–625.

9. Arbinaga F, Fernández□Ozcorta E, Sáenz□López P, Carmona J. The psychological effects of physical exercise: A controlled study of the placebo effect. Scand J Psychol. 2018 Sep 4;59(6):644–52.

10. Helfer SG, Elhai JD, Geers AL. Affect and Exercise: Positive Affective Expectations Can Increase Post-Exercise Mood and Exercise Intentions. ann behav med. 2014 Sep 23;49(2):269–79.

11. Kwan BM, Stevens CJ, Bryan AD. What to expect when you’re exercising: An experimental test of the anticipated affect–exercise relationship. Health Psychol. 2016 Dec 19;36(4):309–19.

12. Turnwald BP, Goyer JP, Boles DZ, Silder A, Delp SL, Crum AJ. Learning one’s genetic risk changes physiology independent of actual genetic risk. Nat Hum Behav. 2018 Nov 29;3(1):48–56.

13. Flowers EP, Freeman P, Gladwell VF. Enhancing the acute psychological benefits of green exercise: An investigation of expectancy effects. Psychology of Sport and Exercise. 2018 Aug 31;39:213–21.

14. Meeuwis S, Middendorp H, Veldhuijzen D, Laarhoven A, Houwer J, Lavrijsen A, et al. Placebo Effects of Open-label Verbal Suggestions on Itch. J Rehabil Med. 2018 Jan 1;98(2):268–74.

15. Locher C, Buergler S, Nascimento AF, Kost L, Blease C, Gaab J. Lay perspectives of the open-label placebo rationale: a qualitative study of participants in an experimental trial. BMJ Open. 2021 Aug 1;11(8):e053346–e053346.

16. Guevarra DA, Moser JS, Wager TD, Kross E. Placebos without deception reduce self-report and neural measures of emotional distress. Nat Commun. 2020 Jul 29;11(1).

17. Finniss DG, Kaptchuk TJ, Miller F, Benedetti F. Biological, clinical, and ethical advances of placebo effects. Lancet. 2010 Feb 1;375(9715):686–95.

18. Ernst E, Saradeth T, Resch KL. The powerful placebo. Lancet. 1991 Mar 1;337(8741):611–611.

19. Lieberman, D. (Eds). Exercised. Cheers Publishing. 2022

20. Kelley JM, Kaptchuk TJ, Cusin C, Lipkin S, Fava M. Open-Label Placebo for Major Depressive Disorder: A Pilot Randomized Controlled Trial. Psychother Psychosom. 2012 Jan 1;81(5):312–4. 21.

21. Carvalho C, Caetano JM, Cunha L, Rebouta P, Kaptchuk TJ, Kirsch I. Open-label placebo treatment in chronic low back pain: a randomized controlled trial. PAIN. 2016 Oct 14;157(12):2766–72.

22. Guevarra DA, Moser JS, Wager TD, Kross E. Placebos without deception reduce self-report and neural measures of emotional distress. Nat Commun. 2020 Jul 29;11(1).

23. Kari-Alyse, L. (2023). Understanding and Harnessing Psychological and Social Forces to Improve Healthcare. Stanford University

24. Saunders B, Saito T, Klosterhoff R, de Oliveira LF, Barreto G, Perim P, et al. “I put it in my head that the supplement would help me”: Open-placebo improves exercise performance in female cyclists. PLoS ONE. 2019 Sep 24;14(9):e0222982.

25. Schneider M, Dunn A, Cooper D. Affect, Exercise, and Physical Activity among Healthy Adolescents. J Sport Exerc Psychol. 2009 Dec 1;31(6):706–23.

26. Schaefer M, Kühnel A, Schweitzer F, Enge S, Gärtner M. Neural underpinnings of open-label placebo effects in emotional distress. Neuropsychopharmacol. 2022 Dec 1;48(3):560–6.

27. Damrongthai C, Kuwamizu R, Suwabe K, Ochi G, Yamazaki Y, Fukuie T, et al. Benefit of human moderate running boosting mood and executive function coinciding with bilateral prefrontal activation. Sci Rep. 2021 Nov 22;11(1).

28. Tempest GD, Eston RG, Parfitt G. Prefrontal Cortex Haemodynamics and Affective Responses during Exercise: A Multi-Channel Near Infrared Spectroscopy Study. PLoS ONE. 2014 May 1;9(5):e95924.

29. Haehl W, Mirifar A, Beckmann J. Regulate to facilitate: A scoping review of prefrontal asymmetry in sport and exercise. Psychol Sport Exerc. 2022 May; 60:102143.

30. Lattari E, Portugal E, Moraes H, Machado S, Santos T, Deslandes A. Acute Effects of Exercise on Mood and EEG Activity in Healthy Young Subjects: A Systematic Review. CNSNDDT. 2014 Jun 12;13(6):972–80.

31. Silveira R, Prado RCR, Brietzke C, Coelho-Júnior HJ, Santos TM, Pires FO, et al. Prefrontal cortex asymmetry and psychological responses to exercise: A systematic review. Physiol Behav. 2019 Jun 17;208:112580–112580.

32. Davidson RJ. Anterior electrophysiological asymmetries, emotion, and depression: Conceptual and methodological conundrums. Psychophysiol. 1998 Sep 1;35(5):607–14.

33. Zhang J, Zhou, RL. Frontal EEG laterality: An index of the capability of emotion regulation. Advance in Psychological Science. 2010,18(11):1679–83.

34. Shephard RJ. PAR-Q, Canadian Home Fitness Test and exercise screening alternatives. Sports Med. 1988 Mar 1;5(3):185–95.

35. Garber CE, Blissmer B, Deschenes MR, Franklin BA, Lamonte MJ, Lee IM, et al. Quantity and Quality of Exercise for Developing and Maintaining Cardiorespiratory, Musculoskeletal, and Neuromotor Fitness in Apparently Healthy Adults. Med Sci Sports Exerc. 2011 Jun 22;43(7):1334–59.

36. Wang YB, Guo L, Fan JY, Mao ZX. Expectations Come True: The Placebo Effect of Exercise on Affective Responses. Res Q Exerc Sport. 2022 Sep 19;94(4):1153–61.

37. Hardy CJ, Rejeski WJ. Not What, but How One Feels: The Measurement of Affect during Exercise. J Sport Exerc Psychol. 1989 Sep 1;11(3):304–17.

38. Kendzierski D, DeCarlo KJ. Physical Activity Enjoyment Scale: Two Validation Studies. J Sport Exerc Psychol. 1991 Mar 1;13(1):50–64.

39. Frijia EM, Billing A, Lloyd-Fox S, Rosas EV, Collins-Jones L, Crespo-Llado MM, et al. Functional imaging of the developing brain with wearable high-density diffuse optical tomography: A new benchmark for infant neuroimaging outside the scanner environment. Neuroimage. 2020 Oct 24;225:117490–117490.

40. Vidal-Rosas EE, Zhao H, Nixon-Hill RW, Smith G, Dunne L, Powell S, et al. Evaluating a new generation of wearable high-density diffuse optical tomography technology via retinotopic mapping of the adult visual cortex. Neurophotonics, 2021. Jun 20; 8(2): 025002.

41. Godin G. The Godin-Shephard leisure-time physical activity questionnaire. The Health and Fitness Journal of Canada, 2011, 4(1): 18–22

42. Hernández-Vicente A, Hernando D, Marín-Puyalto J, Vicente-Rodríguez G, Garatachea N, Pueyo E, et al. Validity of the Polar H7 Heart Rate Sensor for Heart Rate Variability Analysis during Exercise in Different Age, Body Composition and Fitness Level Groups. Sensors. 2021 Jan 29;21(3):902–902.

43. Borgo G. Borg’s perceived exertion and pain scales. Champaign, 1998 IL: Human Kinetics.

44. Bok D, Rakovac M, Foster C. An Examination and Critique of Subjective Methods to Determine Exercise Intensity: The Talk Test, Feeling Scale, and Rating of Perceived Exertion. Sports Med. 2022 May 4;52(9):2085–109.

45. Haas JW, Rief W, Weiß F, Doering BK, Kleinstäuber M, Ruwoldt S, et al. The effect of patient-centered communication on medication intake: an experimental study. Psychology Health & Medicine. 2021 Aug 20;27(10):2138–51.

46. Crum AJ, Salovey P, Achor S. Rethinking stress: The role of mindsets in determining the stress response. J Personal Soc Psychol. 2013 Feb 25;104(4):716–33.

47. Ross KM, Wing RR. “Memory bias” for recall of experiences during initial weight loss is affected by subsequent weight loss outcome. J Behav Med. 2017 Oct 27;41(1):130–7.

48. Ruby MB, Dunn EW, Perrino A, Gillis R, Viel S. The invisible benefits of exercise. Health Psychol. 2011 Jan 1;30(1):67–74.

49. Hargreaves EA, Stych K. Exploring the peak and end rule of past affective episodes within the exercise context. Psychol Sport Exerc. 2012 Oct 11;14(2):169–78.

50. Sagy I, Abres J, Winnick A, Jotkowitz A. Placebos in the era of open□label trials: An update for clinicians. Eur J Clin Invest. 2018 Oct 13;49(1):e13038.

51. Schmitt A, Martin JA, Rojas S, Vafa R, Scheef L, Strüder HK, et al. Effects of low- and high-intensity exercise on emotional face processing: an fMRI face-matching study. Soc Cogn Affect Neurosci. 2019 Jun 1;14(6):657–65.

